# Discovery and validation of plasma-based protein biomarkers for the aetiological distinction of bacterial and non-bacterial febrile infections in African children

**DOI:** 10.1101/2024.01.10.24300882

**Authors:** Jacqueline M Waeni, Timothy K Chege, Elijah T Gicheru, Martin Mutunga, James Njunge, Daniel O’Connor, Charles J Sande

**Author notes:** Correspondence to: Dr. Charles Sande, KEMRI-Wellcome Trust Research Programme, Kilifi, Kenya P.O. Box 230 - 80108.

## Abstract

**Background:** In many low resource settings, the clinical management of children with febrile infections is hindered by poor access to diagnostic tools to determine whether the cause of an infection is bacterial, viral or parasitic. As a result, many clinicians resort to the default prescription of antibiotics as a safety precaution, contributing to the alarming spread of antimicrobial resistance. Commonly used biomarkers for identification of bacterial sepsis such as CRP lack aetiological specificity and are frequently elevated by non-bacterial infections including malaria. We set out to discover and validate new biomarkers for the characterization of the microbial aetiology of febrile acute infections in Kenyan children.

**Methods:** We recruited a discovery cohort comprising of children who had been admitted to hospital with a variety of severe acute infections. Diagnostic identification of viral infections was done using a 15-target virus PCR panel, bacterial infections were identified using blood culture while malaria infections were identified by microscopy. Using mass spectrometry analysis, we identified a set of 76 plasma proteins whose abundance varied significantly by the microbial aetiology of infection and used machine learning to generate a shortlist of candidate biomarkers that had the highest diagnostic performance in distinguishing aetiologies. To validate these candidate biomarkers, we recruited a separate validation cohort where the plasma levels of the shortlisted biomarkers were assayed among children with different infectious aetiologies using a custom protein microarray.

**Results:** In the discovery study, six candidate biomarkers whose plasma abundance was significantly different in children with bacterial and viral infections were shortlisted by random forest for cross-cohort validation (AGT, HRG, LBP, PON1, SERPINA1, SERPINA3). In the validation study, we found that of the six biomarkers, only AGT compared favourably to CRP and identified febrile bacterial infections with a sensitivity of 72.4% (95% CI 48.4% - 83.6%) compared to CRP which distinguished febrile bacterial infections with a sensitivity of 69.5% (30.8% - 88.2%). Plasma AGT was superior to CRP in distinguishing children with febrile bacterial infections from those with febrile malaria episodes, with a sensitivity of 72.5% (40% - 84.6%) for AGT and 26% (15% - 32.8%) for CRP.

**Conclusions:** We report the discovery of AGT, as a sensitive plasma biomarker for the identification of febrile bacterial infections among African children living in a malaria endemic setting.

## Introduction

The care of children with potentially life-threatening febrile acute infections in the world’s poorest countries remains a challenging problem due to the inadequate access to pathogen diagnostics in these settings.(*1*) Understanding the microbial aetiology underlying febrile acute paediatric infections is important in devising effective clinical management strategies, including decisions on whether to initiate antibiotic therapy. In much of sub-Saharan Africa, the clinical diagnosis of severe acute paediatric infections is based on the WHO’s Integrated Management of Childhood Illnesses (IMCI), a set of clinical guidelines to support the identification and management of the most common paediatric causes of severe disease and death(*2*). These guidelines are however largely based on the assessment of clinical symptoms at presentation and provide limited sensitivity for establishing whether the cause of an infection is viral, bacterial or parasitic. As a consequence of this imprecision, the IMCI guidelines recommend the empiric administration of antibiotics to most severely ill children as a contingency measure in absence of objective diagnostic data to rule out non-bacterial causes of infection. The resulting excess administration of antibiotics to large numbers of children without proven bacterial infection is now thought to be a major driver for the development of antimicrobial resistance and is an increasingly ominous threat to global health.(*3*) Low cost point of care (PoC) diagnostic biomarkers designed to distinguish bacterial infections from other infectious causes of acute illness in children have been evaluated extensively in high resource settings and the most widely used of these, c-reactive protein or CRP, is generally elevated in children with invasive bacterial infections.(*4–7*) The main drawback against the use of CRP as a diagnostic PoC test for establishing the microbial aetiology of paediatric infections is the fact that it is a non-specific marker of systemic inflammation that can be elevated by a wide range of triggers including traumatic injury(*8*) and parasitic infections such as plasmodium falciparum.(*9*) This lack of aetiological specificity limits the utility of CRP as a diagnostic PoC test for bacterial infections in many parts Africa and Asia, where parasitic infections such as falciparum and vivax malaria are endemic.(*10, 11*) To address the shortcomings inherent in the use of non-specific acute phase protein biomarkers such as CRP, a number of studies have used transcriptome analysis to identify host genes that are differentially expressed during viral and bacterial infection and that could therefore be leveraged as aetiological biomarkers for these infections. Early data from these efforts has been promising - for example a large biomarker discovery study in European children reported a two transcript gene expression signature that can classify bacterial and viral infections with a sensitivity and specificity >90%.(*12, 13*) Many other studies have confirmed these associations and have reported equally encouraging diagnostic performance for a range of alternative transcriptional biomarkers.(*14*) Despite the clear promise demonstrated by these novel transcriptome-based approaches it is unlikely that these technologies will achieve widespread adoption in resource poor settings - the cost of implementing complex, highly technical and resource-intensive tests for routine use in resource-deprived settings is likely to represent a considerable and possibly insurmountable access barrier. In these settings, the most feasible technologies for routine diagnostic use need to be based on low cost technologies that are easy to execute without the requirement of additional laboratory resources or technology-intensive equipment. Most low cost PoC tests, including lateral flow tests (LFT), are designed to detect proteins in blood and other body fluids(*15*) and almost all PoC diagnostic applications in low resource settings - ranging from pregnancy testing(*16–18*) to rapid malaria diagnostics(*19*) - are based on the detection of host or pathogen proteins in different body fluids. Therefore, to maximise the likelihood of adoption, biomarkers that are designed for the aetiological distinction of febrile acute infections in these environments need to be protein-based.

In order to identify new protein biomarkers for differentiating febrile acute bacterial infections from other acute infections, we designed a study to discover and validate new aetiology-specific plasma protein biomarkers in Kenyan children. To achieve this, we recruited a discovery cohort and used mass spectrometry proteomics to quantify plasma proteins from children with microbiologically- or PCR-proven bacterial and viral infections. We then used machine learning to generate a ranked shortlist of candidate biomarkers, whose abundance in plasma varied by microbial aetiology. To validate these candidate biomarkers, we recruited a separate validation cohort in which their relative abundance in plasma was assayed using protein microarrays and their diagnostic performance compared to CRP.

## Results

The overall design of this study is illustrated in **Fig. 1**. In the biomarker discovery phase of the study, plasma samples from 63 children who were admitted with microbiologically proven bacterial infections and 75 who were admitted with PCR proven viral infections were included in the analysis (**Fig. 2 & Table 1**). The identities of the individual viral and bacterial species that were studied in the cohort are shown in **Fig. 3a**. Using high performance liquid chromatography tandem mass spectrometry (HPLC-MS/MS), we quantified a total of 418 proteins in the plasma of these children - 26 of these proteins were significantly higher in children with bacterial infections while 50 were significantly higher in children with viral infections (**Fig 3b)**. Selected examples of plasma proteins that were differentially expressed in the two aetiological groups are shown in **Fig 3c**. We then used the machine learning aggregator SIMON (*20*) to rank the diagnostic performance of all the differentially expressed proteins in distinguishing infections of bacterial origin from those of viral origin. Of the 91 machine learning models that were tested on SIMON, the regularised random forest model was the best performing model with an AUC of 0.9 on the training data set and an AUC of 0.93 on the test data set. We applied this model on all plasma proteins identified in the discovery study and used the variable importance function to shortlist the top performing proteins based on feature importance. The top seven protein predictors of infectious aetiology were angiotensinogen (AGT), Serpin Family A Member 1 (SERPINA1), Serpin Family A Member 3 (SERPINA3), Paraoxonase 1 (PON1), Histidine Rich Glycoprotein (HRG/B2R8I2), lipopolysaccharide binding protein (LBP) and CRP (**Fig. 3e**). A second random forest model was trained that included only the top biomarkers whose variable importance score in the initial model was > 50%. The performance of this seven biomarker model was assessed in a training and test data set. The top 7 biomarkers chosen were AGT, SERPINA1, SERPINA3, PON1, B2R8I2 (HRG), LBP and CRP. **Fig 3f** shows AUC curves obtained when the top 7- biomarker model was used to predict aetiology.

**Fig. 1.**
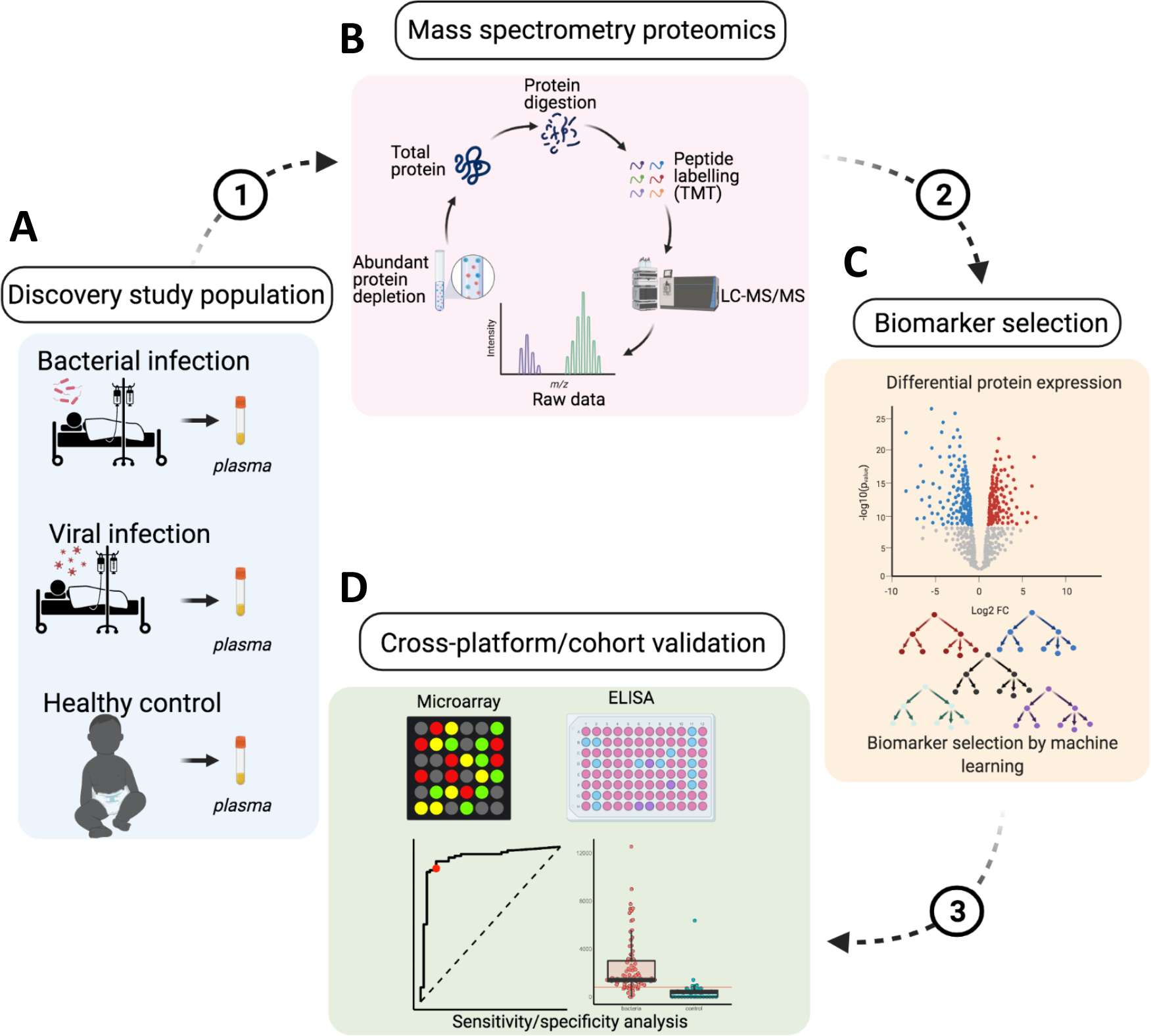

**Fig. 2.**
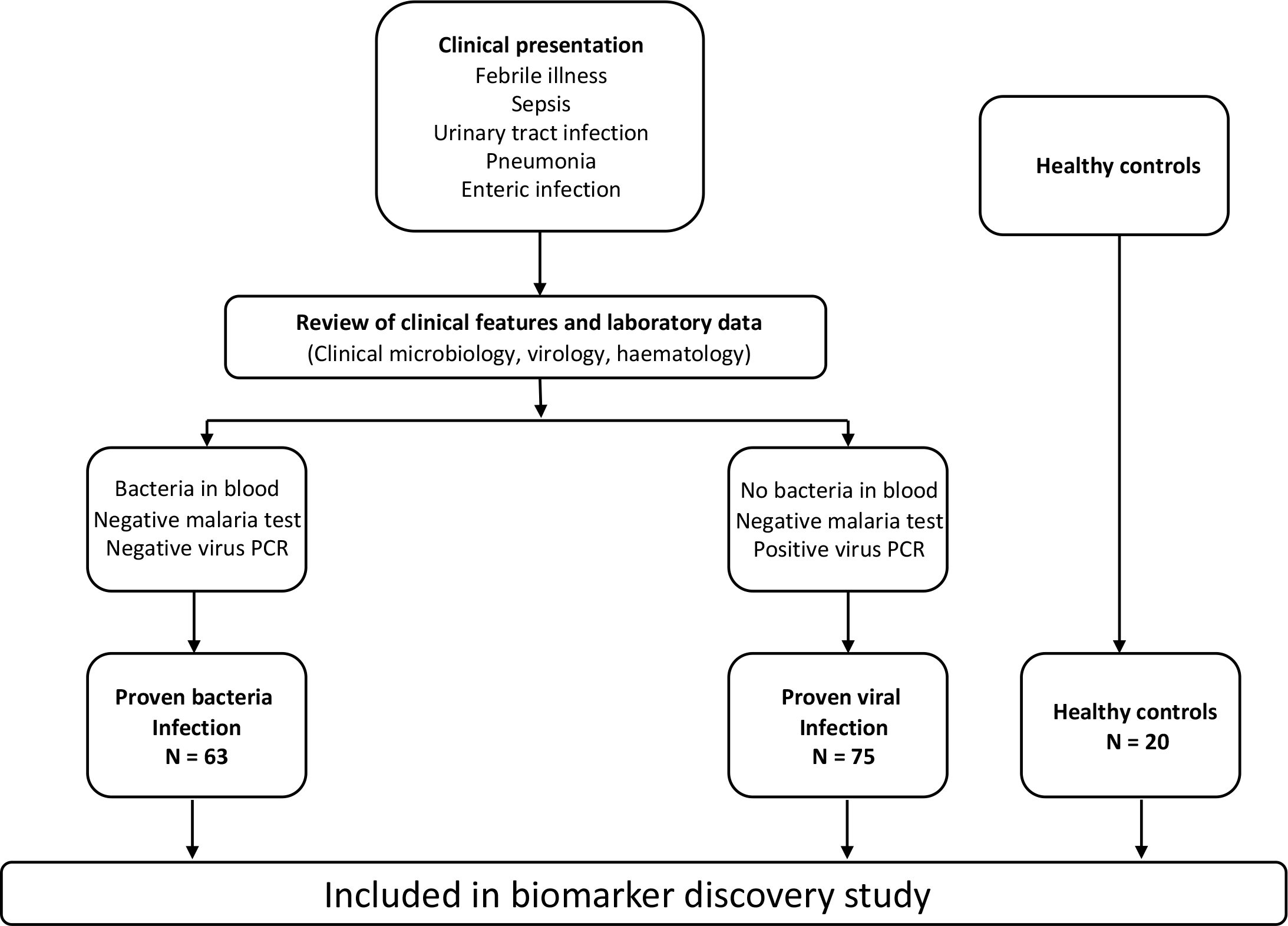

**Fig. 3.**
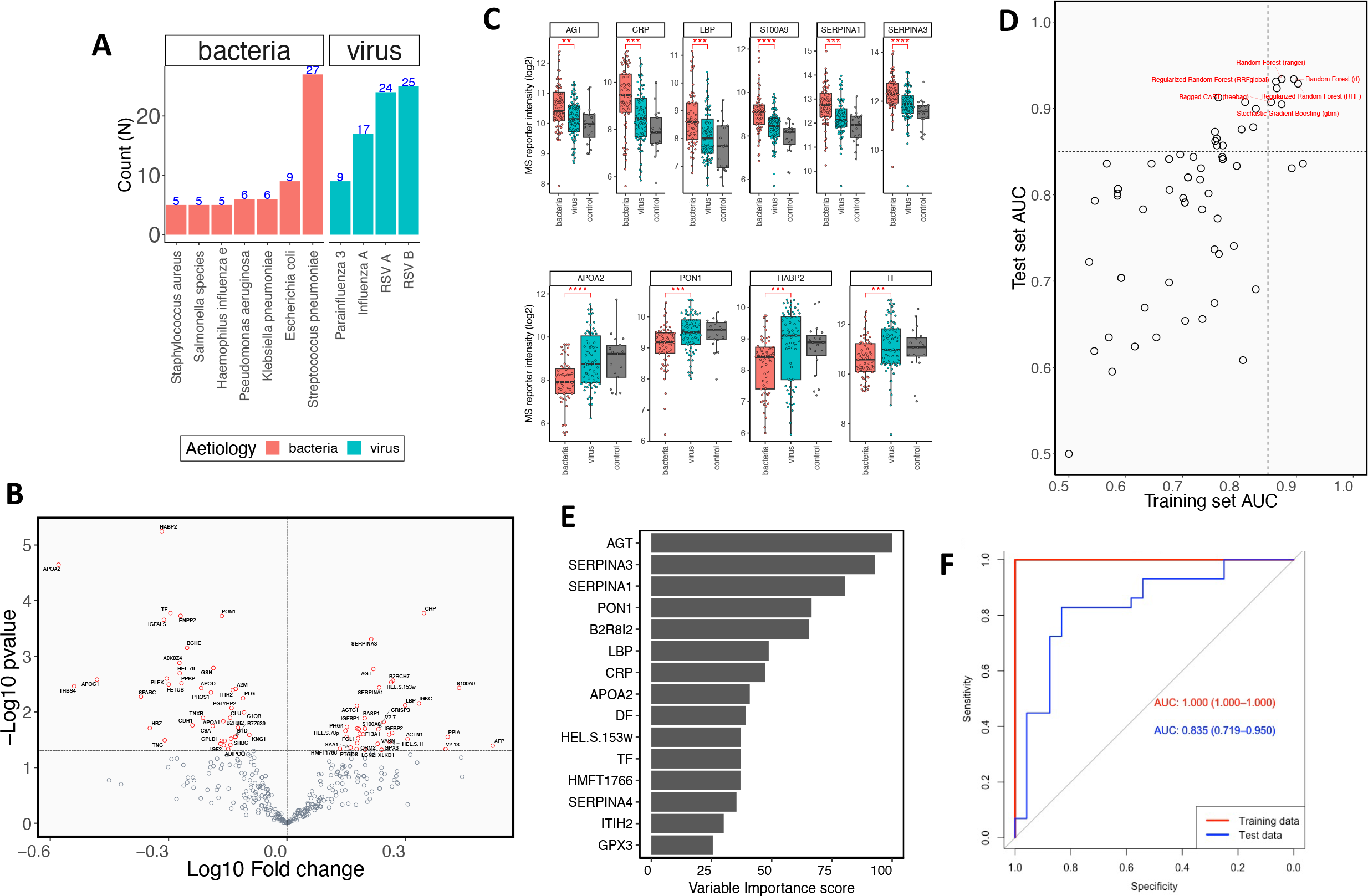

**Table 1.**

| Characteristic | Virus,<br>N = 75<sup>1</sup> | Bacteria,<br>N = 63<sup>1</sup> |
| --- | --- | --- |
| Gender (Female %) | 39% | 39% |
| Age (Months) | 10 (3, 18) | 7 (2, 13) |
| **Clinical Features at presentation** |  |  |
| Mid upper arm circumference (cm) | 13.0 (12.0, 14.0) | 11.4 (9.8, 13.2) |
| Weight-for-height Z-score | -0.9 (-1.8, -0.1) | -1.4 (-1.9, -0.8) |
| Length-for-age Z-score | -0.9 (-1.8, 0.2) | -1.6 (-2.5, -0.8) |
| Respiratory rate (BPM) | 59 (47, 67) | 62 (52, 71) |
| Oxygen saturation (%) | 95 (92, 98) | 94 (87, 97) |
| Axillary Temperature (°C) | 38.2 (37.5, 39.0) | 38.7 (37.8, 39.6) |
| Cough, No. (%) | 73 (97%) | 43 (75%) |
| Breathing Difficulty, No. (%) | 72 (96%) | 54 (95%) |
| Lower chest wall indrawing, No. (%) | 73 (97%) | 51 (89%) |
| Flaring, No. (%) | 42 (56%) | 37 (65%) |
| Hypoxia, No. (%) | 12 (16%) | 18 (32%) |
| Fever, No. (%) | 57 (76%) | 45 (79%) |
| Diarrhoea, No. (%) | 10 (13%) | 11 (19%) |
| Vomiting, No. (%) | 14 (19%) | 11 (19%) |
| Leucocytosis, No. (%) | 23 (34%) | 24 (59%) |
| Deaths, No. (%) | 3 (4.0%) | 19 (33%) |
| **Full Blood Count** |  |  |
| Haemoglobin (g/dl) | 9.4 (8.4, 10.9) | 8.1 (6.9, 10.1) |
| Platelets x10<sup>3</sup>/ml (range) | 394 (300, 526) | 341 (226, 514) |
| White Blood Cells (x10<sup>3</sup>/ml) | 11 (9, 16) | 18 (7, 26) |
| Red Blood Cells (x10<sup>6</sup>/l) | 4.4 (4.0, 4.9) | 3.6 (3.1, 4.5) |
| Lymphocytes (10<sup>3</sup>/ml) | 4.0 (3.2, 6.3) | 4.3 (2.5, 6.4) |
| Neutrophils (10<sup>3</sup>/ml) | 4.2 (2.8, 6.4) | 4.3 (0.9, 11.7) |
| Monocytes (10<sup>3</sup>/ml) | 1.5 (0.9, 2.5) | 2.7 (1.5, 6.0) |
| <sup>1</sup>Median (IQR) or Frequency (%) |  |  |

To validate the seven candidate biomarkers that were identified in the discovery study we recruited an independent validation cohort of children under the age of five years who were admitted to hospital with different acute infections. The cohort was made up of 212 children who had proven bacterial infections and 239 children who had proven viral infections. 76 children had indeterminate infections (i.e. virus, bacteria and malaria co-infections), 202 had unknown infections (negative virus PCR, negative blood culture and negative malaria blood slide), 39 were diagnosed with malaria parasites, while 29 children were age-matched healthy controls. The diagnostic flowchart that was used to establish these aetiological definitions is shown in **Fig. 4** and the clinical and demographic characteristics of children in the cohort are shown in **Table 2**. The most commonly isolated bacterial species were *Staphylococcus aureus*, *Escherichia coli*, *Streptococcus pneumoniae* and *Salmonella species* while the most commonly identified viral pathogens were RSV, Human metapneumovirus, Parainfluenza 3 and Influenza A as shown in **Fig 5a**. The plasma levels of six of the seven biomarkers identified in the discovery study (AGT, HRG, LBP, PON1, SERPINA1, SERPINA3) were quantified using a customised protein microarray whose design is illustrated in **Fig 5b** while the levels of the seventh biomarker, CRP was measured using the ILab Aries clinical biochemistry analyser. Of the 6 biomarkers tested on the microarray, only AGT exhibited greater sensitivity in the detection of bacterial infections compared to CRP (**Fig 5g**). A cohort-wide analysis of AGT and CRP showed that the plasma levels of both biomarkers were significantly higher in children with proven bacterial infections compared to those with proven viral infections as well as healthy controls (**Supplementary figure 2**). In a subset analysis of febrile children who had axillary temperatures ≥ 38°C at the time of admission, we found that at a threshold of 12 log2 MFI, plasma AGT could distinguish febrile bacterial infections from febrile viral infections with a sensitivity of 72.4% (95% CI 48.4% - 83.6%) and a specificity of 76.9% (95% CI 54.9% - 85.2%) – **Fig 5d**, while at an optimised threshold concentration of 82.2mg/L the sensitivity of CRP in detecting bacterial infections among febrile children was 69.5% (95% CI 30.8% - 88.2%) with a specificity of 92.9% (95% CI 50% - 100%) – **Fig 5f**. We then assessed the independent effect of falciparum malaria on plasma AGT and CRP in order to define the aetiological specificity of these biomarkers. There was no difference in the concentration of plasma CRP between febrile children with invasive bacterial infections and those infected with plasmodium falciparum parasites (p=0.19, **Fig. 5i**). In contrast, we found that plasma AGT in febrile children with bacterial infections was significantly higher than those of febrile children infected with plasmodium falciparum (p<0.0001, **Fig 5i)**. At an optimized threshold of 12 log2 MFI plasma AGT could distinguish febrile children with bacterial infections from those infected with plasmodium falciparum parasites with a sensitivity of 72.5% (95%CI 40% - 84.6%). For CRP, an optimised threshold of 99.7 mg/L, provided a sensitivity of 26% (95% CI 15% - 32.8%) for distinguishing between febrile bacterial infections and febrile malaria infections (**Fig 5j)**.

**Fig. 4.**
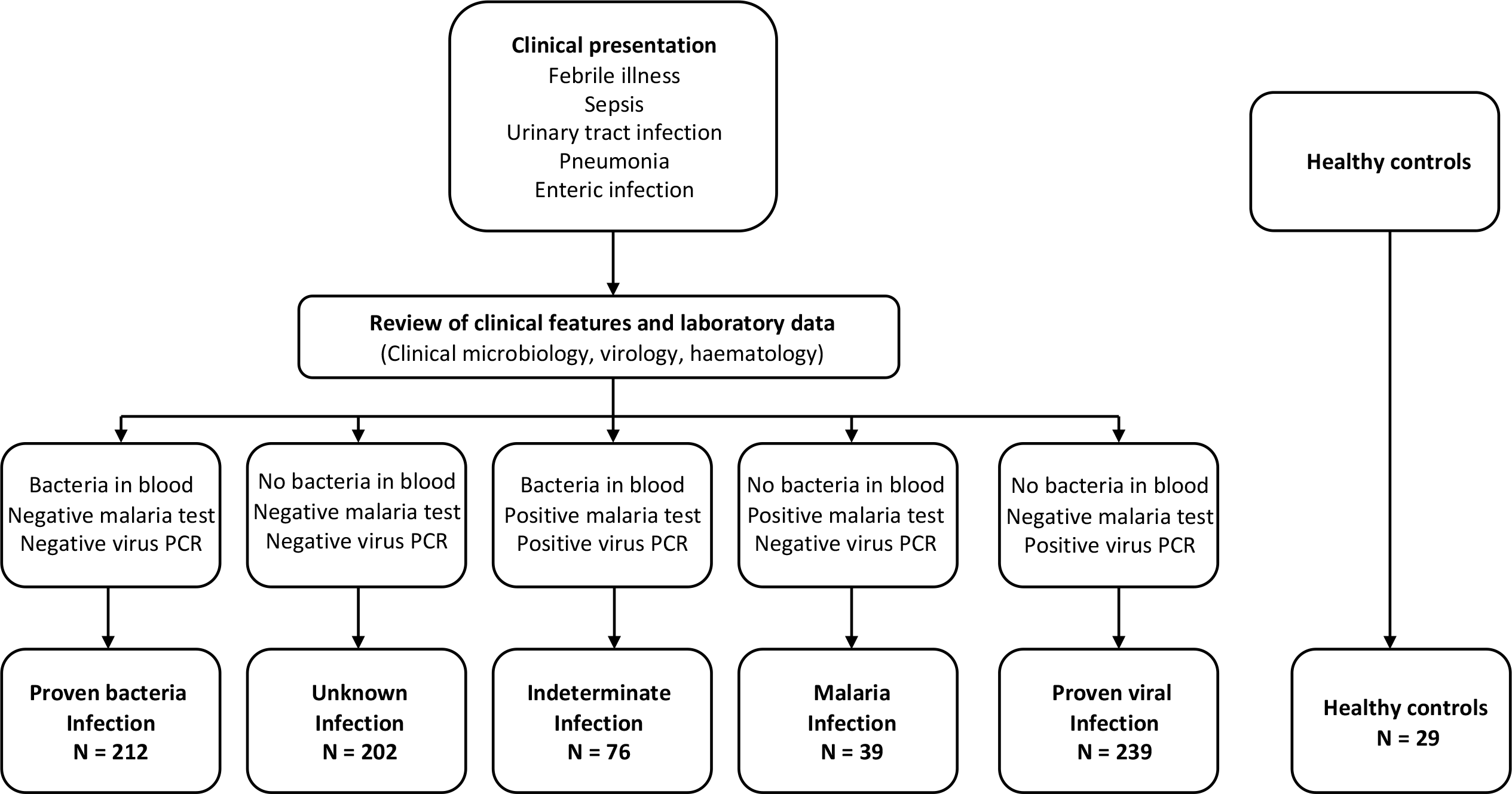

**Fig. 5.**
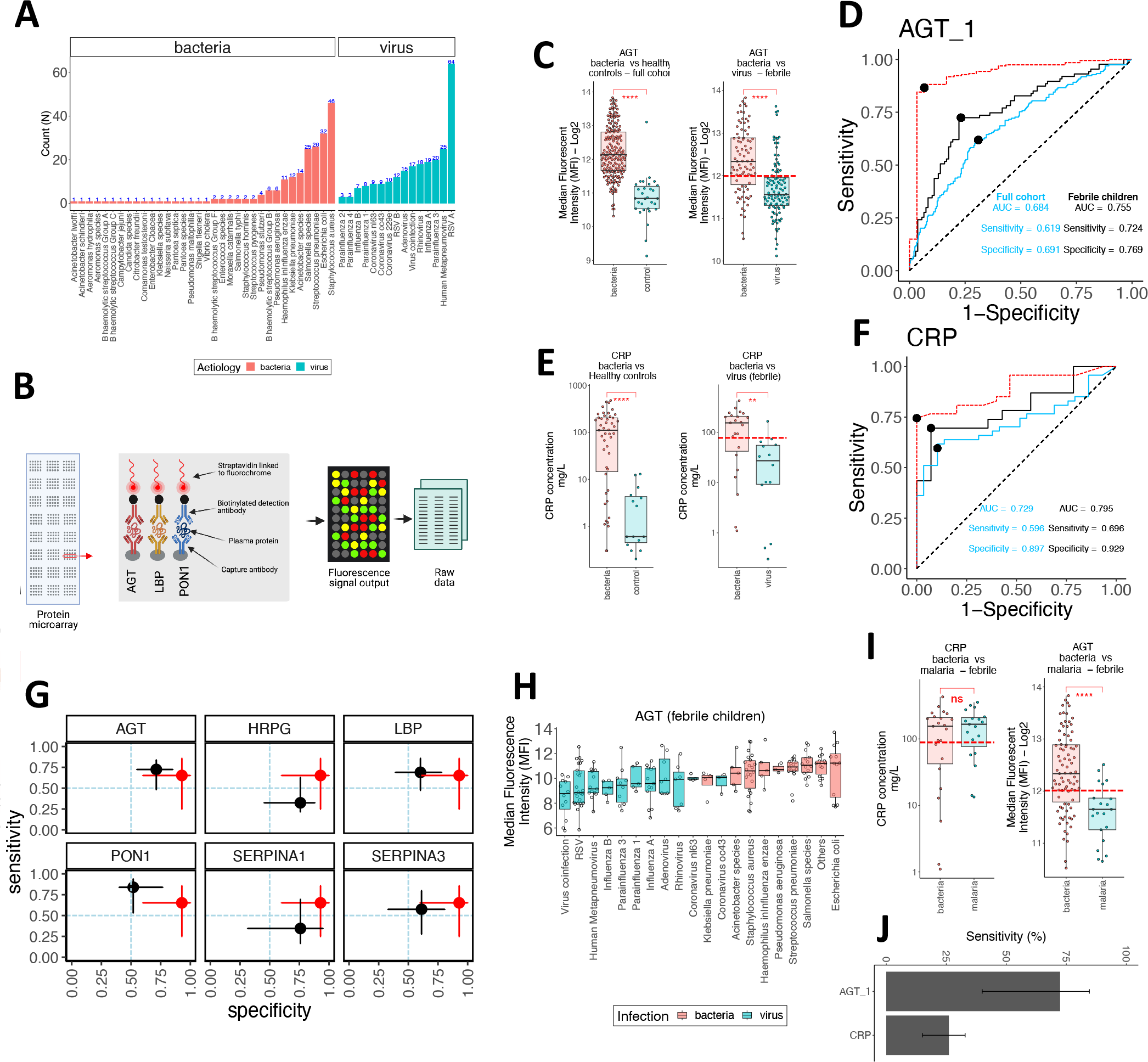

**Table 2.**
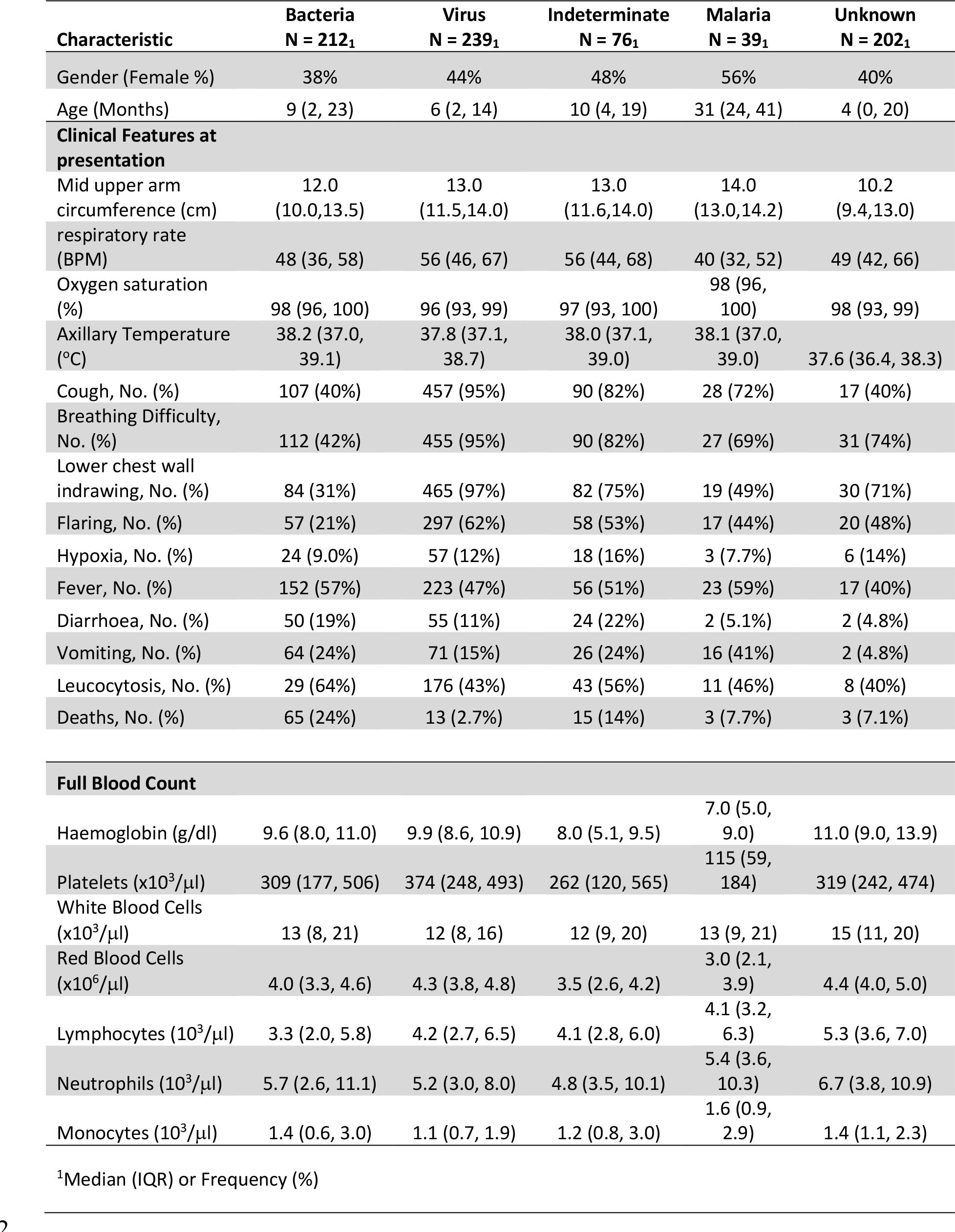

To visualize the differences in pathogen-specific abundance of plasma AGT in febrile children, we calculated the median AGT level on the basis of the infecting pathogen and ranked these median abundances in ascending order. With the exception of children infected with *Klebsiella pneumoniae,* the median abundance of AGT in children infected with all other bacterial species was higher than the median plasma abundance of AGT in all virus-infected children, regardless of the species of the infecting virus (**Fig. 5h**). To understand whether the application of AGT as an aetiological biomarker for bacterial infections could potentially inform clinical decisions regarding the initiation of antibiotic therapy, we compared antibiotic prescription rates between febrile children who had been admitted with confirmed viral infections and those who had unknown infections (negative blood culture, negative virus PCR and negative malaria blood slide). While there was no statistical difference in the plasma AGT levels of children with febrile viral infections compared to febrile children with unknown aetiologies (**Fig. 6a**) only 25% of children with proven viral infections had an antibiotic prescription at discharge(**Fig. 6b**), compared to 55% of children in the unknown infection category who were given antibiotics at discharge(**Fig. 6c**).

**Fig. 6.**
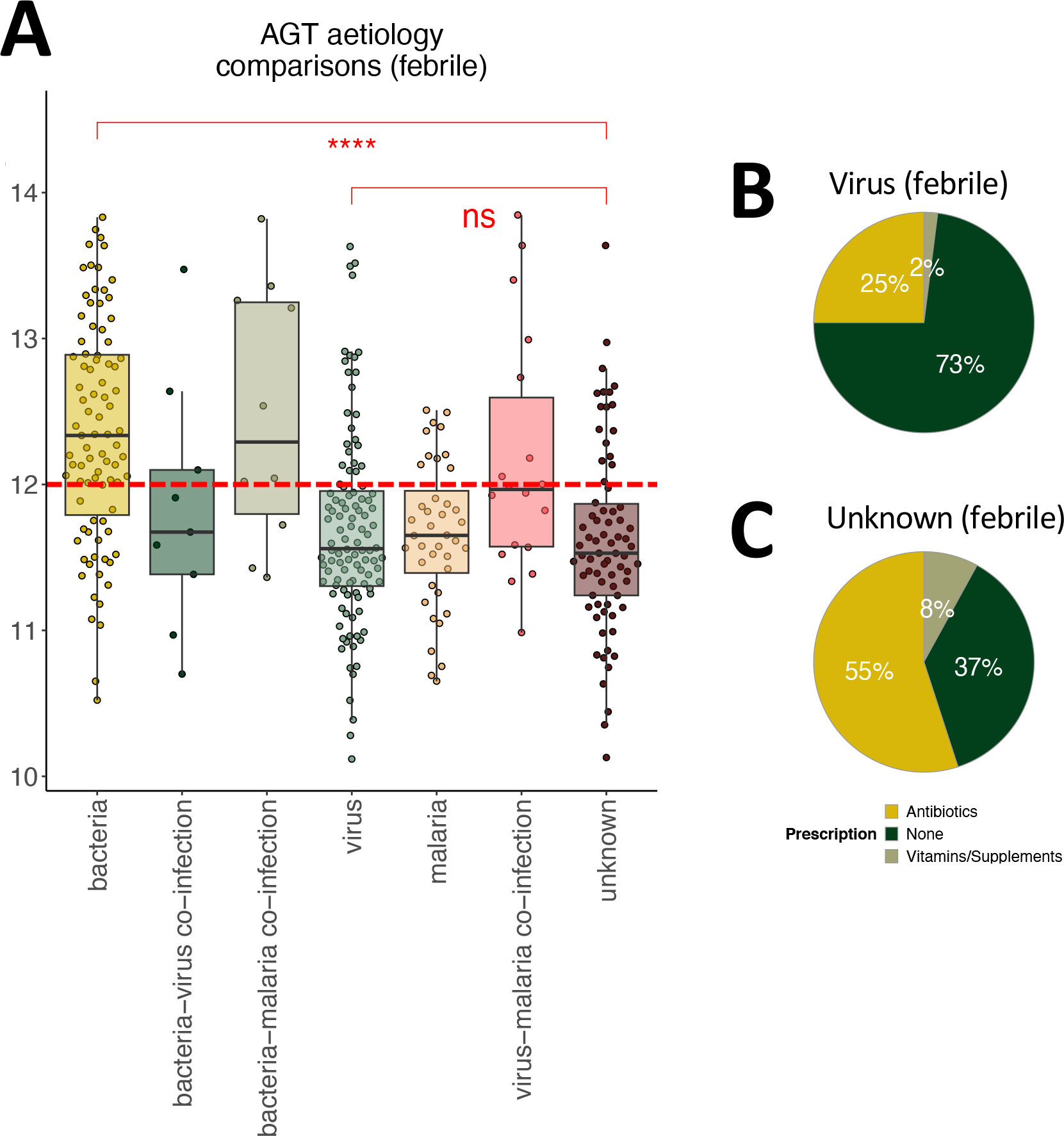

## Discussion

The aim of the present study was to discover and validate new plasma protein biomarkers that could be used to distinguish febrile acute illnesses caused by bacterial infections from other infectious causes of febrile illness in children. We report the discovery of AGT as a sensitive and specific plasma protein biomarker for the identification of bacterial infections in the plasma of febrile children from a low income setting in Kenya. The performance of AGT compared favorably to CRP in distinguishing febrile infections caused by bacteria from those caused by viruses and was superior to CRP in distinguishing invasive bacterial infections from febrile malaria episodes.

In order to maximize the likelihood of identifying new biomarkers of infection, we used HPLC M/S M/S, a high throughput and ultra-sensitive proteomics platform(*21, 22*) in the discovery phase of the study. This decision was informed by the fact that many of the current generation of infection biomarkers that are used in clinical practice, are based on serendipitous discoveries that were made many decades ago using technologies that were available at the time. CRP, for example, was discovered almost 100 years ago as a protein that was highly abundant in the sera of febrile patients with invasive pneumococcal disease, that reacted with the with the carbohydrate antigen in the pneumococcus capsule.(*23*) Another commonly used biomarker for febrile bacterial illness, Procalcitonin (PCT), was discovered over three decades ago as a marker of bacterial sepsis following previous observations of increased serum calcitonin in patients with a range of non-communicable inflammatory disorders.(*24*) In the decades that followed these discoveries, powerful new tools for the unbiased screening of the human plasma proteome were developed and refined, including HPLC MS/MS. These technologies allow for the untargeted measurement of the full human plasma proteome,(*25*) thereby allowing for the identification of proteins whose abundance in plasma is selectively altered by different biological conditions or infectious stimuli. We took advantage of these technological advancements to conduct a large-scale, untargeted proteomics screen using plasma obtained from children with microbiologically- and PCR-proven bacterial and virus infections. We identified >400 proteins in plasma of these children and used machine learning to shortlist six biomarkers with the highest diagnostic performance in distinguishing bacterial infections from viral infections. We then proceeded to validate these candidate biomarkers in a different cohort of children, using the protein microarray technology. The decision to validate the shortlisted biomarkers using a separate cohort and protein detection technology was predicated on the observation that many recent discoveries made using omics technologies have been not successfully replicated in subsequent studies. This failure of reproducibility has been attributed to many factors ranging from differences in the detection technologies are used in discovery studies compared to those that are used in replication studies to nuanced differences in study populations that impact the likelihood of concordance.(*26, 27*) To unequivocally establish the veracity of the shortlisted biomarkers, we recruited a separate cohort of children who had been admitted with different acute infections and designed a new immunoassay-based protein microarray to quantify the abundance of the biomarkers in plasma. Using this cross-cohort and cross-platform approach, we found that only one of the six candidate biomarkers retained its superior diagnostic performance relative to CRP – plasma AGT was more sensitive than CRP in distinguishing febrile bacterial infections from febrile viral infections and crucially, it was also more sensitive in distinguishing febrile bacterial infections from febrile malaria episodes. The latter distinction is an especially critical requirement for biomarkers that are intended for use in settings with a high malaria burden. Previous studies of children in malaria-endemic settings in Africa and South East Asia have shown that while plasma CRP is generally present at low levels in febrile children with viral infections, it is significantly elevated in children with febrile malaria episodes and invasive bacteremia, making it impossible to differentiate the latter two aetiologies on the basis of plasma CRP alone.(*28*) Findings from the present study indicate that plasma AGT is specific to the bacterial aetiology, and that it can be used to identify bacterial infections with high sensitivity even against the background of malaria endemicity. This aetiological specificity suggests that in these settings AGT may be a superior biomarker for guiding care decisions on antibiotic prescription compared to CRP and PCT - which is also elevated during severe febrile malaria infections.(*29, 30*) However more studies in diverse populations with different patterns of disease endemicity are needed to rigorously evaluate the effectiveness of AGT under such conditions.

To model the potential impact of AGT on clinical decisions regarding the initiation of antibiotic therapy to febrile children, we compared the AGT expression profiles of febrile children with established aetilogies to those of febrile children for whom the underlying cause of infection could not be established. We reasoned that the latter population of children with unknown aetiologies would mirror most febrile children presenting to health facilities in resource limited settings with no capacity to identify the aetiology of infection using conventional diagnostic tools. While the abundance of AGT in the plasma of children with unknown aetiologies was significantly lower than in children with bacterial infections, it was essentially identical to those with viral infections. Despite the broad similarity in AGT expression between children with viral and unknown aetiologies, more than 55% of the children with unknown aetiologies were given an antibiotic compared to 25% of the children with a known viral infection. This represents a greater than 100% excess in antibiotic administration to children in the unknown aetiology group who were unlikely to be having bacterial infections and who, therefore, would not have benefitted from antibiotic therapy. A possible explanation for the excess prescription of antibiotics to children with undifferentiated fevers is that while the majority of febrile illnesses in childhood are self-resolving viral infections,(*31*) in the absence of objective diagnostic data to rule out bacterial infections, presumptive antibiotic treatment to all febrile children for whom a definitive microbiological diagnosis has not been established is often the only recourse left to clinicians in low resource settings.(*32*) The immediate clinical imperative to minimize risk to the relatively small proportion of febrile children who may have life-threatening invasive bacterial disease likely outweighs broader considerations for more targeted antibiotic therapy with the view to limiting the global spread of antimicrobial resistance. The findings in this study suggest that the use of plasma AGT as a diagnostic biomarker to guide clinical decisions on antibiotic therapy, might mitigate against antibiotic overuse in many low income settings.

## Methods

### Study site, population and procedures

This study was conducted at the Kilifi County Hospital, a paediatric referral facility on the Coast of Kenya, serving a predominantly rural, agrarian community. Children under the age of 5 years presenting to hospital with a range of severe acute infections were recruited at admission and a sample of blood was collected for bacterial culture, full blood count, separation of plasma and identification of malaria parasites by microscopy. In addition to blood samples, nasopharyngeal and oropharyngeal swabs were collected and used for multiplex PCR diagnosis of 15 common childhood viral infections, including RSV (A & B), parainfluenza virus (1, 2, 3 and 4), adenovirus, influenza (A, B and C), coronavirus (OC43, NL63 and e229), human metapneumovirus and rhinovirus. A standardised set of clinical and anthropometric measurements were collected from all study participants including age, sex, axillary temperature, among others. Data obtained from clinical and microbiological investigations was used to classify children into different aetiologies as follows: children were deemed to have a proven invasive bacterial infection if bacteria were isolated from blood, none of the targets on the viral PCR panel returned a positive result and no malaria parasites were isolated from blood. Children were deemed to have a proven viral infection if they had positive results on the virus PCR panel, had a negative blood culture results and a negative malaria blood slide. Children who had negative blood cultures, negative virus PCR results and a negative malaria blood slide were classified as having unknown aetiologies while those whose diagnostic results indicated co-infections with bacteria, viruses and/or with malaria parasites were classified as have indeterminate aetiologies – these functional definitions are shown in detail in **Fig 2**. A control group comprising of healthy, age-matched children was recruited from home and a blood sample obtained for comparison. This study was conducted in accordance with Good Clinical Laboratory Practise (GCLP) principles and was approved by the Kenya Medical Research Institute Scientific Ethics Research Unit.

### Biomarker discovery - study design and analysis

For the discovery study, only samples from children with proven viral or bacterial aetiologies as well as the healthy controls were utilized for biomarker discovery experiments. Discovery of aetiology-specific biomarkers in the discovery cohort was carried out using high performance liquid chromatography tandem mass spectrometry (HPLC MS/MS). To improve sensitivity for detection of low-abundance plasma proteins, the top twelve most abundant proteins in plasma were first depleted using depletion spin columns. The depleted proteins were: α1-Acid Glycoprotein, fibrinogen, α1-Antitrypsin, haptoglobin, α2-Macroglubulin, IgA, albumin, IgG, apolipoprotein A-I, IgM, Apolipoprotein A-II and transferrin (Thermo Scientific). Preparation of depleted plasma samples for mass spectrometry analysis was conducted as previously described.(*33*) Briefly, the protein concentration of each sample was measured using the Bradford assay and was subsequently standardized to 15 µg of total protein for each sample. Protein reduction was done by adding 40 mM Dithiothreitol (DTT) to each sample after which alkylation was undertaken by adding 80 mM Iodoacetamide (IAA) to each sample. 400 µl ice-cold acetone was then added to the samples, which were then vortexed and incubated for 1 hour at -20°C to precipitate the proteins. Protein digestion was then undertaken using 1 µg trypsin per sample. TMT Mass Tag Labelling kits (Thermo Scientific) were used to label sample batches to facilitate multiplexing. 13 μl of the TMT labeling reagent was added to each 100 μl sample, after which equal amounts of each sample were pooled in a new microcentrifuge tube and concentrated by Speedvac to reduce peptide sample volume to 10 μl. Peptide purification was then done using ZipTip pipette tips and quantified using the Qubit protein assay. Peptides were then loaded using an ultra-high-pressure liquid chromatography system and chromatographic separation of peptides carried out on a reverse-phase column over a 180-min elution gradient. Peptides were measured using liquid chromatography instrumentation consisting of a Dionex Ultimate 3000 nano-flow ultra-high-pressure liquid chromatography system coupled via a nano-electrospray ion source to a Q Exactive Orbitrap mass spectrometer (Thermo Scientific). Raw mass spectrometer files were analysed by MaxQuant(*34*) version 1.6.0.131 by searching against the human Uniprot FASTA database (downloaded December 2018) using the Andromeda search engine. The false discovery rate (FDR) was set to 0.01 for both proteins and peptides and a maximum of two missed cleavages were allowed in the database search. A minimum of two unique peptides for a protein were considered a positive identification. The maxquant output file was exported to R (version 4.2.3) for analysis. A detailed description of the mass spectrometry experiments is included in Supplementary methods.

### Biomarker validation - study design and analysis

To validate the candidate biomarkers identified in the discovery study, we recruited a separate cohort of children admitted to KCH with severe acute infections. The recruitment criteria was identical to the discovery cohort and the same set of clinical, anthropometric and diagnostic data were collected. We then developed a customised protein microarray chip to simultaneously quantify the plasma levels of candidate biomarkers that were shortlisted in the discovery study. Microarray slides were produced by depositing onto epoxy-coated polished glass slides, two 100 picolitre droplets containing 250ng/ml of capture antibodies that were specific to defined epitopes present on candidate biomarker proteins. Microarray production was done by non-contact printing using a commercial microarrayer (Aj003S Arrayjet Sprint). Each slide was divided into 24 identical mini arrays onto which a full set of capture antibodies, each targeting a different candidate biomarker protein, was identically printed in triplicate. The slides were then dried at ambient temperature and scanned on a fluorescent scanner (Genepix 4300A) to obtain background slide fluorescence that would subsequently be subtracted from the final array data set to adjust for non-specific fluorescence. Slides were then loaded onto hybridization cassettes, and washed three times with phosphate buffered saline (PBS). The slides were then incubated with a blocking buffer comprising PBS containing Tween-20 and 5% bovine serum albumin (BSA) for an hour at 37°C. A one in thirty dilution of human sera was prepared using PBS-tween-20 & 5% bovine serum albumin, added into different mini arrays and incubated at room temperature for three hours. Biotinylated antibodies targeting alternative epitopes on the candidate biomarker proteins were then added to each mini array and incubated for two hours at room temperature after which the slides were washed as previously indicated. Streptavidin linked to Alexafluor 647 was then added at a dilution of 1:800 and incubated for 2 hours. The slides were then washed thrice with PBS-Tween 20, rinsed with Milli-Q water and then analysed on a Genepix 4300A fluorescence scanner. The resulting data exported to R software for analysis. To facilitate comparison between biomarkers in validation study and CRP, we measured the levels of plasma CRP in a subset of children using the ILab Aries clinical biochemistry analyser.

### Data analysis

To identify plasma proteins that were differentially expressed by aetiology in the discovery cohort, linear regression was performed with an empirical bayes model on normalized mass spectrometry reporter intensity values from children with bacterial or viral infections using the ‘limma’ package in R. A Benjamini and Hochberg (BH) false discovery rate (FDR) alpha value of 0.05 was used as the threshold for differential protein expression between groups. To identify plasma proteins that could distinguish children with bacterial infection from those with viral infection, the machine learning model aggregator, SIMON was used.(*20*) SIMON (Sequential Iterative Modelling “OverNight”) is a free and open-source software that enables the building and evaluation of more than 200 machine learning algorithms based on the ‘caret’ library in R. The discovery cohort dataset was split into two as follows: 60% of the data set wad randomly partitioned as the training set and 40% as the test or validation set for SIMON analysis. Next, classification models were trained using 183 machine learning algorithms which were applied iteratively on the training set. Area under the Receiver operating characteristic curves (AUCROC) were used to assess the performance of the machine learning classification models. Models with the highest combined AUC on the training and test data sets were considered to have the best predictive performance. Other measures of model performance such as sensitivity, specificity and accuracy were used to filter out poorly performing models. In the validation study, microarray mean fluorescence intensity (MFI) values were compared between viral and bacterial aetiologies for each candidate biomarker that was shortlisted from the discovery study. We used the ‘cutpointr’ package in R to determine optimal cutpoints that were applied on MFI data to distinguish between aetiologies. These cutpoints were used to calculate the sensitivity and specificity of different biomarkers in the validation study. Detailed analysis of the microarray data set described in supplementary methods.

## Data Availability

All data produced in the present work are contained in the manuscript

## Author statement

The authors of this paper have no conflicts of interest to declare. None of the authors or their respective institutions received any payments or services in the past 36 months from a third party that could be perceived to influence, or give the appearance of potentially influencing, the submitted work. This work was funded by the Wellcome Trust (Ref: 105882/Z/14/Z). The authors and their institutions have not received payment or services from a third party for any aspect of the submitted work.

## SUPPLEMENTARY MATERIALS

### Supplementary methods

#### HPLC MS/MS plasma proteomics

Identification and quantification of proteins in the plasma of children in the discovery cohort was carried out using high performance liquid chromatography tandem mass spectrometry. Ten microlitres of plasma was added directly to the resin slurry in the column and incubated for 60 minutes at room temperature after thorough mixing. The column was then centrifuged for 2 minutes at 1000 x g and the filtrate was used to determine protein concentration using the Bradford assay. 250 μl of Bradford reagent was added into each well of an ELISA and 5 μl of bovine serum albumin (BSA) standards of known concentration were added in duplicate into respective wells, followed by test samples. Optical density was read at an absorbance of 595nm using a microtitre plate reader after incubating the plates for 10 minutes at room temperature. A standard curve was generated using the manufacturer supplied standards and used to calculate the concentration of study samples. Sample concentrations were then used to determine sample volumes required to achieve 15 µg of total protein for each sample. The sample volume was adjusted to 100 µl by adding appropriate volumes of 100 mM Triethylammonium bicarbonate (TEAB). Protein reduction was done by adding 40 mM Dithiothreitol (DTT) to each sample, followed by a one hour incubation at 70°C with shaking. Alkylation was done by adding 80 mM Iodoacetamide (IAA) to each sample followed by incubation in the dark at room temperature for 1 hour. 400 µl ice-cold acetone was then added samples, which were then vortexed and incubated for 1 hour at -20°C to precipitate the proteins. The samples were centrifuged for 10 minutes at 15000 x g and supernatants were discarded without dislodging the protein pellet. The acetone was left to evaporate at room temperature for 30 minutes and protein was resuspended in 100mM TEAB with thorough vortexing to dissolve the protein pellet. Trypsin digestion was done using 1 µg of trypsin per sample followed by 16 hour incubation at 37°C on a shaking platform. Thermo Scientific TMT Mass Tag Labelling kits was used to label sample batches to facilitate multiplexing. TMT labelling reagents were equilibrated to room temperature by centrifuging the vials briefly for 1 minute at 15000 x g. 42μl of anhydrous acetonitrile was added to each TMT tube and allowed to dissolve for 5 minutes with occasional vortexing. 13 μl of the TMT labeling reagent was added to each 100 μl sample. 8μl of 5% hydroxylamine was then added to the sample and incubated for 15 minutes at room temperature to quench the reaction. Equal amounts of each sample were pooled in a new microcentrifuge tube and concentrated by Speedvac to reduce peptide sample volume to 10 μl. Peptide purification was then done using ZipTip pipette tips which contained C18 reversed-phase media for desalting and concentrating peptides. Purified peptide concentration was determined using the Qubit protein assay. Peptides (1 µg) were loaded using a Dionex Ultimate 3000 nano-flow ultra-high-pressure liquid chromatography system (Thermo Scientific) on to a 75µm x 2cm C18 trap column (Thermo Scientific). Chromatographic separation of peptides was carried out on a reverse-phase 25cm-long column (Thermo Scientific) maintained at 40°C over a 180-min elution gradient (2 to 30% of mobile phase B; 80% acetonitrile with 0.1% formic acid) at a flow rate of 0.3μl/min. Peptides were measured using LC instrumentation consisting of a Dionex Ultimate 3000 nano-flow ultra-high-pressure liquid chromatography system (Thermo Scientific) coupled via a nano-electrospray ion source (Thermo Scientific) to a Q Exactive Orbitrap mass spectrometer (Thermo Scientific). The msg^1 settings were; Resolution, 70000; Automatic gain control (AGC) target, 3e6; maximum injection time, 100ms; scan range, 380- 1600m/z; while the ms^2 settings were: Resolution, 17500; AGC target, 5e4; maximum injection time, 100ms; isolation window, 1.6 m/z. The top 10 most intense ions were selected for ms^2 and fragmented with higher-energy collision fragmentation using normalized collision energy of 28 and these ions were subsequently excluded for the next 20s.

#### Analysis of plasma protein levels using custom protein arrays

Visualization of variance in median fluorescence intensity values before and after quality control was carried out in order to identify outliers and non-biological sources of variation in the data. This was done using boxplots and principal components analysis. As shown in **Suppl fig. 1a**, before outlier filtration and normalization, there was substantial technical variation observed in the data. Principal components analysis of the raw data showed the first principal component (PC) was responsible for 52.6% of the variation while the second PC was responsible for 19% variation. Samples formed 6 main clusters based on the first and second PC. This clustering is attributable to batch effects from microarray slides and different days of sample processing **Suppl fig. 1c**. Principal variance components analysis (PVCA) of the data showed that prior to QC, the leading drivers of variation were different batches in which samples were processed. Samples that were processed in 2020 were variable from those that were processed in 2021 and from PVCA results in **Suppl fig. 1c**, this explained 50% of the variation in that data. Processing samples on different days also explained 10% of the variation as well slide to slide variation that explained 15% variation. Therefore, due to evidence of technical variation within the data, normalization and batch correction steps were applied. Correction for technical variance was performed on the data using Combat algorithm in the ‘sva’ package in R and Principal components analysis was done after batch correction to evaluate the impact of the correction. The first PC was associated with 48% of the variation in the data while PC2 accounted for 16% of the variation. Clustering by year and day of sample processing was no longer visible **Suppl fig. 1d**. Analysis of the sources of variation using PVCA on batch corrected data revealed minimal variation explained by technical artefacts such as analytical batching, year and day of sample processing **Suppl fig. 1f**. Residual variation, representing biological variance in the data increased from 25% to 98% after correction for technical variance **Suppl fig. 1f**.

---

**Supplementary figure 1.**
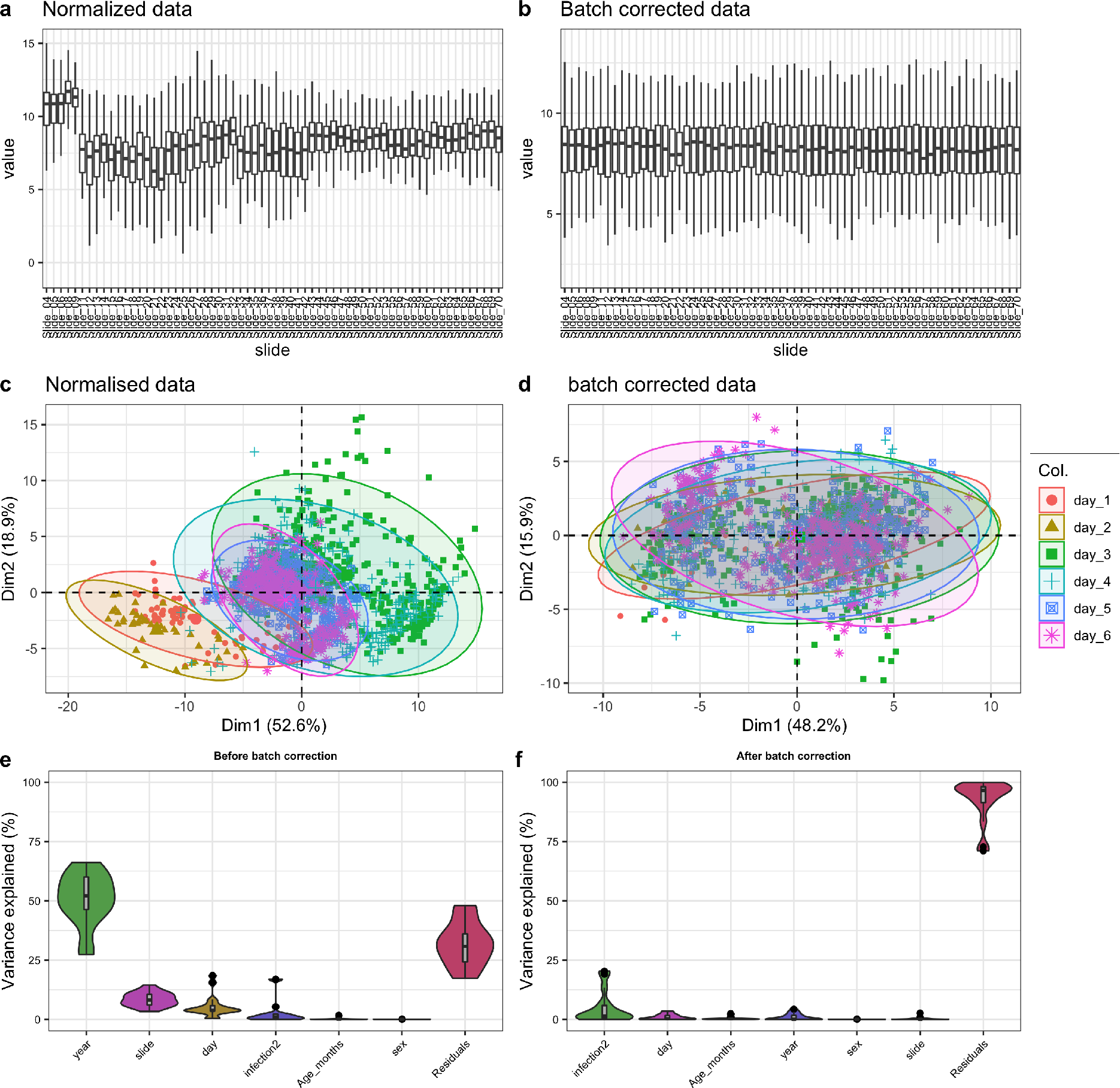
a) Boxplots of raw intensities for each slide included in the analysis before QC. b) Boxplots of batch corrected intensities c) PCA of raw intensities showing clustering according to the day of sample processing d) PCA of batch corrected intensities showing no clustering by day of sample processing after batch correction e) Violin plots showing amount of variance in the data explained by different batch variables before batch correction and f) after batch correction. Before QC, most of the variation in the data is attributable to technical sources such as batching by year, day and slide. This effect is reduced after batch correction and residual variation in the data is magnified.

**Supplementary figure 2.**
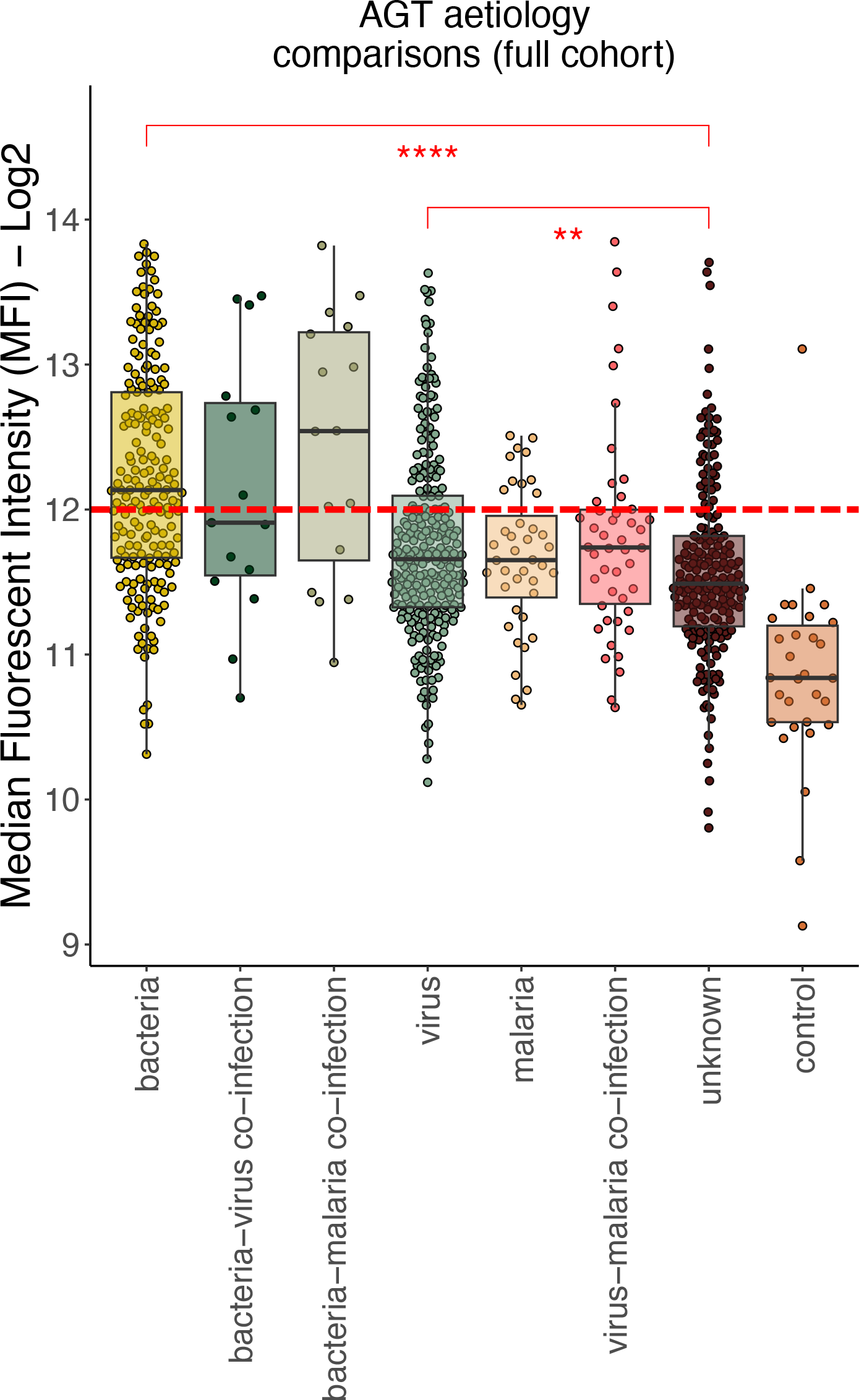
Aetiological comparisons of plasma AGT expression across the entire validation cohort

